# Effectiveness of COVID-19 vaccines against SARS-CoV-2 infection and symptomatic infection in the England Winter Coronavirus (COVID-19) Infection Study cohort

**DOI:** 10.1101/2024.12.30.24319550

**Authors:** Nurin Abdul Aziz, Nick Andrews, Christopher Rawlinson, Andrew Buckley, Alexander Allen

**Affiliations:** Immunisations and Vaccine-Preventable Diseases Division, UK Health Security Agency; Surveillance Strategy and Transformation Branch, UK Health Security Agency

## Abstract

**Background:** The Winter Coronavirus (COVID-19) Infection Study (WCIS) was a sample-based household study in England and Scotland aiming to inform COVID-19-related epidemiology and health pressures over the 2023-2024 winter period. We aim to estimate vaccine effectiveness (VE) against both SARS-CoV-2 infection in general and specifically symptomatic infection (COVID-19) within the WCIS cohort in England.

**Methods:** Data from self-reported lateral flow device (LFD) tests for SARS-CoV-2 were collected from participants alongside self-completed questionnaires from November 2023 to March 2024. A test-negative case-control design was used to estimate VE of the Autumn 2023 COVID-19 boosters against SARS-CoV-2 infection and symptomatic infection compared with being unboosted, regardless of previous vaccination history. Symptomatic infection as an outcome was restricted to participants who reported an ARI symptom associated with their test. Logistic regression was used to calculate VE, with LFD test result as the outcome, vaccination status as primary exposure variable, and adjustment for covariates. Analyses were also stratified by bivalent BA4/5 and monovalent XBB vaccines to assess VE of individual vaccine types.

**Results:** The analysis included 109,929 English residents primarily in the 60-80 age range (54%), of White ethnic background (95%), and in the least deprived quintile (31%). Participants contributed 114,066 eligible tests in the analysis, of which 42,340 were associated with a symptomatic outcome. VE against infection peaked at 49% (95% Confidence Interval (CI): 35-60%) at 2-4 weeks post-vaccination, with waning to a null effect occurring after 10 weeks (VE: 5% (95% CI: -5-14%)). Similarly, VE against symptomatic infection peaked at 49% (95% CI: 32-63%) after 2-4 weeks, waning after 10 weeks (VE: 5% (95% CI: -7-16%)). The bivalent vaccine showed low and mainly non-significant evidence of protection against either outcome, whereas the monovalent vaccine showed a peak VE of 49% (95% CI: 34-60%) at 2-4 weeks against infection and 49% (95% CI: 31-63%) at 2-4 weeks against symptomatic infection.

**Conclusions:** The Autumn 2023 COVID-19 vaccine campaign provided moderate protection against infection and symptomatic infection during the 2023 winter period, with a differential effect between vaccine type. Household studies such as WCIS are useful to understand impacts of vaccination campaigns within the community, especially in the post-pandemic period where testing capacity is restricted to hospital settings.

## Background

COVID-19, caused by the SARS-CoV-2 virus, has evolved since its emergence, causing waves of infection and severe disease (1–3). COVID-19 vaccination has been shown to provide protection against hospitalisation and other severe outcomes throughout the changing disease profile of COVID-19 (4–6). A primary course of COVID-19 vaccine was initially rolled out to all adults in England in June 2021, followed by a booster dose launched in November 2021 (7). Since then, routine programmes occurring each spring and autumn have been established to protect the groups most vulnerable to severe disease, based on age and clinical risk.

In England, vaccine effectiveness (VE) against symptomatic infection could also be assessed due to the availability of widespread community testing prior to 1 April 2022 (8– 10). In more recent periods, the large majority of testing is available only in healthcare settings, limiting surveillance, including VE analyses, to hospitalised cases. (11).

To better understand COVID-19 activity within the community, and inform healthcare pressures over the winter period attributable to the virus, the Winter Coronavirus (COVID-19) Infection Study (WCIS) was launched from November 2023 to March 2024 in England and Scotland to gather epidemiological information on COVID-19.This followed the autumn 2023 COVID-19 vaccination programme, which was launched in September 2023 (12). This vaccination programme targeted those aged 65 and over, older care home residents, those in a clinical risk group aged 6 months or over, and health and social care staff. The programme initially offered a bivalent BA4/5 vaccine, after which a monovalent XBB vaccine also became available a few weeks after.

In this study, we aimed to use the data from the WCIS in England to estimate effectiveness and quantify any waning of the autumn 2023 vaccines against COVID-19 infection and symptomatic infection.

## Methods

### WCIS study design and population

The WCIS was launched as a collaboration between the UK Health Security Agency (UKHSA) and the Office for National Statistics and ran from 13 November 2023 to 7 March 2024 (13). The study included about 150,000 participants from England and Scotland aged 3 years and above. The study population was sampled from participants of the Coronavirus (COVID-19) Infection Survey (CIS), the previous iteration of the WCIS, who consented to be approached for other research studies and took part in the digital version of the CIS (14). No financial incentives were given for participation in the study. The initial CIS study was based on a random sample of households to provide a nationally representative survey.

Participants were asked to test for SARS-CoV-2 using lateral flow devices (LFDs) and report their results through an online questionnaire within a 7-day testing window once every 4-5 weeks. In the event of a positive test, participants were asked to test every other day until two consecutive negative tests were reported. A set of 14 LFDs were posted by mail to participants at the start of the study, and no additional LFDs were sent if all tests were used prior to the end of the study.

The questionnaire was used to collect data on SARS-CoV-2 infection status, dates of SARS-CoV-2 tests, onset dates of symptoms consistent with COVID-19 for symptomatic individuals, health and social care worker status, and information on age, sex, ethnicity, postcode, and region. Postcode information from the WCIS was used to determine the deprivation status of participants using the Index of Multiple Deprivation (IMD) quintile, an area-based measure of relative deprivation.

The analysis population included all WCIS participants living in England who submitted at least one survey between 13 November 2023 and 7 March 2024 and had an available NHS number for linkage to the Immunisation Information System (IIS).

### Linked data sources

#### Immunisation Information System

The UKHSA Immunisation Information System (IIS) contains data on vaccination history and additional demographic characteristics of the population of England registered with a GP (15). WCIS data was linked to the IIS using NHS number. Vaccination data from the IIS was preferentially used over the self-reported vaccination information derived from the WCIS questionnaire. Data on clinical risk group status at the start of the autumn 2023 programme, provided by NHS Cohorting as a Service (CaaS), was extracted from the IIS data (16).

#### Second-Generation Surveillance System

Participants in the WCIS data were also linked using NHS number to the Second-Generation Surveillance System (SGSS), the national laboratory reporting system in England that captures routine laboratory data on infectious diseases, including positive SARS-CoV-2 specimens (17). Data on positive COVID-19 episodes were used to determine if participants had any documented evidence of COVID-19 infection prior to the study or if positive tests captured near the start of the study may have been ongoing from an infection prior to the study. This also allowed assessment of the effect of adjustment for prior infection in the model.

### Statistical methods

Vaccine effectiveness was calculated using a nested test-negative case-control (TNCC) design within the cohort. This approach was selected to be similar to the test-negative approach used for community vaccine effectiveness assessment when community testing was common. The exposure variable for primary analyses was vaccination status in autumn 2023, with SARS-CoV-2 infection (positive vs negative) and symptomatic infection (positive vs negative) as separate end points. Multivariable logistic regression was used and VE was calculated as 1-OR with 95% confidence intervals. For the infection end point, cases were defined as positive SARS-CoV-2 tests from WCIS participants and controls were negative SARS-CoV-2 tests from WCIS participants. For the symptomatic infection end point, cases were positive SARS-CoV-2 tests from WCIS participants with acute respiratory illness (ARI)-associated symptoms, and controls were negative SARS-CoV-2 tests from participants the same ARI-associated symptoms. ARI-associated symptoms included fever, cough, shortness of breath, loss of smell or taste, and noisy breathing or wheezing in under 5-year-olds.

To select the positive and negative tests only the earliest submitted test completed was selected per testing window for each individual. From this, we excluded negative tests taken within 7 days of a previous negative test and negative tests up to 7 days before a positive test. Any tests, positive or negative, within 90 days after a positive test were also excluded. This followed the same criteria as previous used for TNCC studies (8). The final step was to select one random negative test per participant in the study period and, where applicable, the first positive test in the study period per participant. This final step was replicated for the subgroup of tests associated with ARI symptoms for the symptomatic infection endpoint. All analyses included adjustment for age at the start of the autumn 2023 campaign (five-year bands), test week, clinical risk group status coded as those with no evidence of clinical risk, those with a clinical risk other than immunosuppression, and those who are severely immunosuppressed as defined in the Green Book (7), NHS region, index of multiple deprivation (IMD) decile, ethnicity, and healthcare or social care worker status. Influenza vaccination status was also included for adjustment, defined as vaccination with an influenza vaccine in the 2023/2024 season at least 14 days before. To account for the effect of recent COVID-19 infections prior to the study, we adjust for evidence of any COVID-19 infections occurring from a year before the first submitted questionnaire up to 90 days before. Individuals with missing NHS number, age, clinical risk status, test week, region, and ethnicity were removed from the analysis; this comprised 3.5% of the original study population.

An individual was considered vaccinated if, at the time of their test, they had received the autumn 2023 booster 84 or more days after their previous dose, and the vaccine received was the bivalent BA 4/5 vaccine or the monovalent XBB vaccine administered between 1 September 2023 to 14 April 2024. The comparator group was all individuals who did not receive an autumn 2023 vaccine at the time of their test regardless of previous vaccination history. To assess waning, vaccination status was stratified by time to SARS-CoV-2 test after vaccination in the autumn 2023 programme at the following intervals: less than 2 weeks, 2 to 4 weeks, 5 to 9 weeks, and 10 or more weeks. Tests that occurred 0 to 2 days after vaccination were removed to account for the healthy vaccinee bias, where those exhibiting early symptoms of COVID-19 and of poorer health than controls are less likely to seek vaccination. VE was estimated overall and separately for the bivalent BA4/5 and monovalent XBB vaccines. Sensitivity analyses were done restricting to those who had had at least two previous vaccine doses, removing negative tests from individuals who had contributed a positive test, removing positive tests from individuals where the episode may have begun prior to the study based on linkage to prior test data, and removing adjustment for prior infections.

## Results

A total of 518,491 tests were collected from the WCIS from 139,453 participants from England and Scotland. For the nested TNCC study, there was a total of 114,066 eligible tests from 109,929 unique participants in England. There were 108,252 controls and 5,814 cases in the infection outcome. There were 42,340 eligible tests for the symptomatic infection outcome, including 38,288 controls and 4,052 cases (Figure 1). The majority of the study population are over 60 years old, of white ethnicity, and reside in less deprived areas (IMD decile over 6). Full descriptive characteristics are available in Supplementary Table 1.

**Figure 1.**
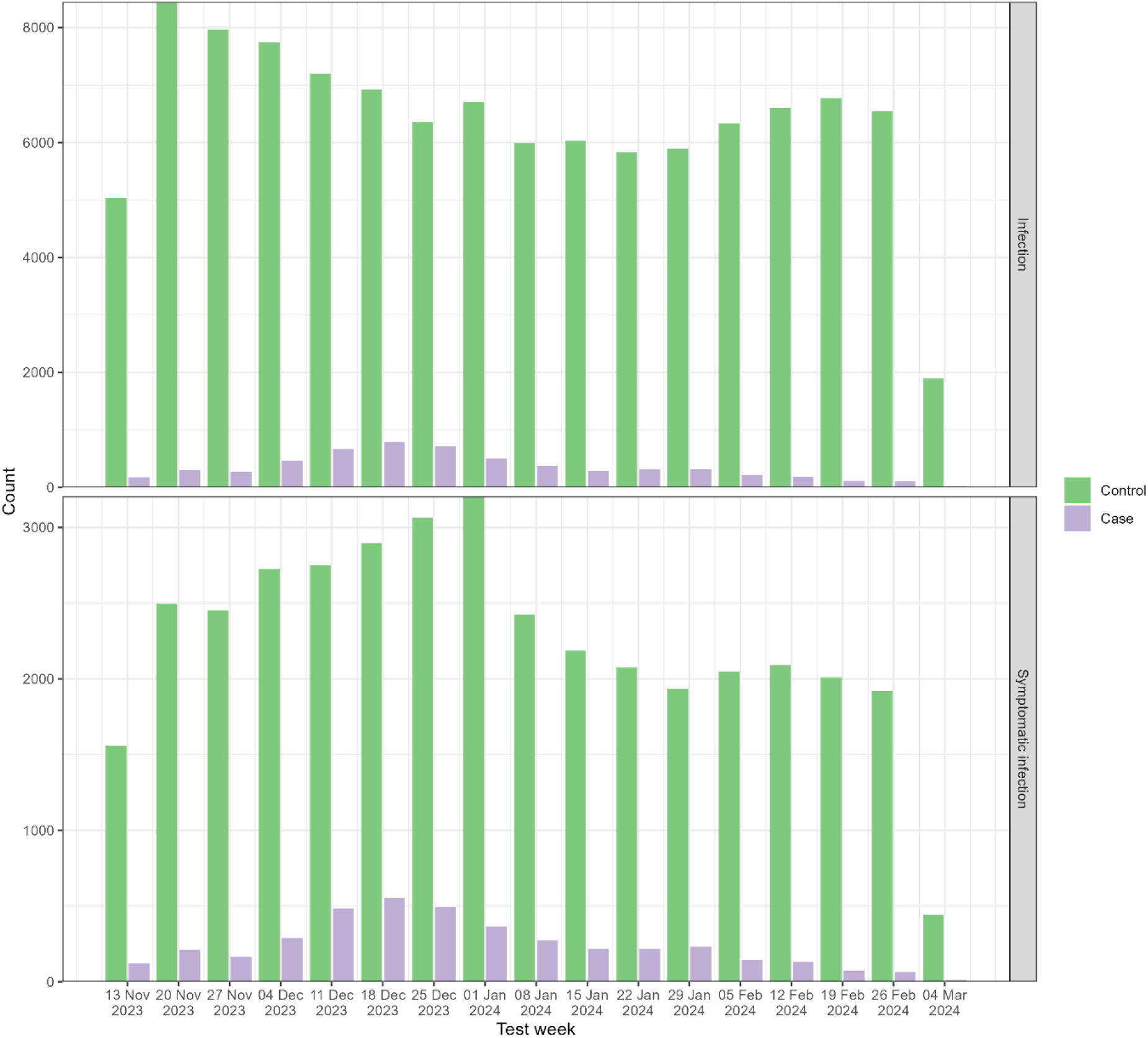
Distribution of cases and controls by week of test and outcome, 13 November 2023 – 7 March 2024.

Figure 2 details the distribution of tests by time to test since vaccination, stratified by vaccine type. Due to the staggered rollout of the bivalent BA4/5 and monovalent XBB vaccines, earlier time intervals of 7 weeks or less since vaccination was mainly represented by those who received the monovalent XBB vaccine.

**Figure 2.**
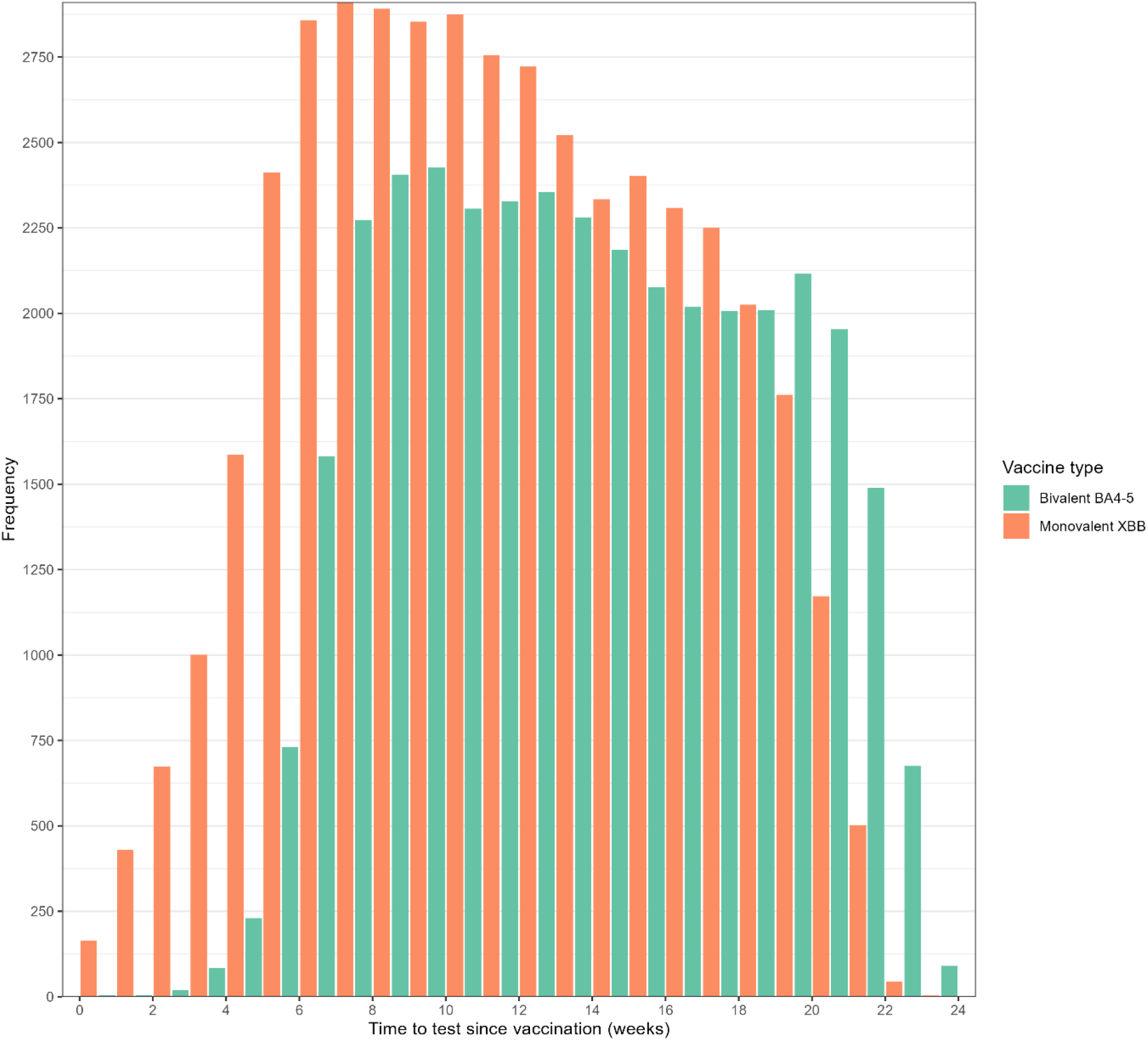
Distribution of time to WCIS test (weeks) since vaccination by vaccine type, 13 November 2023 – 7 March 2024

### Effectiveness of Autumn 2023 vaccine programme

There is evidence that the autumn 2023 programme provided significant protection against SARS-CoV-2 infection in the Winter CIS cohort (Figure 3). VE peaked at 49% (95% CI: 35-60%) 2 to 4 weeks after vaccination, followed by waning to 5% (95% CI: -5-14%) by 10 to 14 weeks.

**Figure 3.**
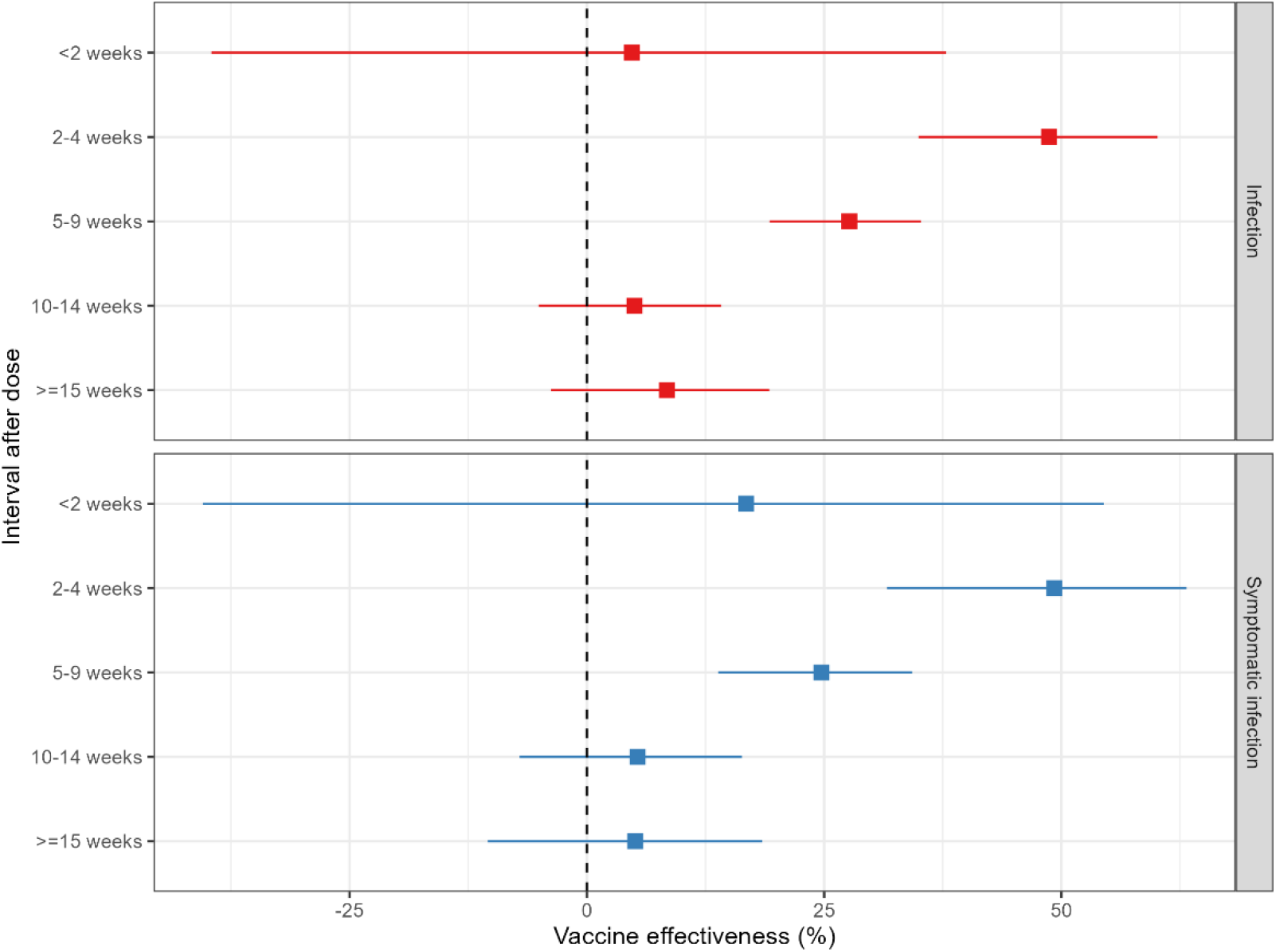
Vaccine effectiveness (VE) against infection and symptomatic infection amongst the WCIS study population, stratified by time since vaccination. Error bars represent 95% confidence intervals. Data behind estimates shown are presented in Supplementary Table S2.

When stratified by vaccine type (Figure 4), the monovalent XBB vaccine showed VE peaking at 49% (95% CI: 34-60%) at 2 to 4 weeks, decreasing to 9% (95% CI: -7-23%) by 15 or more weeks. For the bivalent BA4/5 vaccine, there were very few tests taken <5 weeks from vaccination. The vaccine showed statistically significant VE against infection at 16% (95% CI: 0.3-29%) 5-9 weeks after vaccination, with other intervals showing non-significant VE with wide confidence intervals. To test whether VE differed between vaccines a likelihood ratio test was performed to compare models with combined and separate vaccine effects by manufacturer. This gave a p-value of 0.02 indicating evidence of a difference.

**Figure 4.**
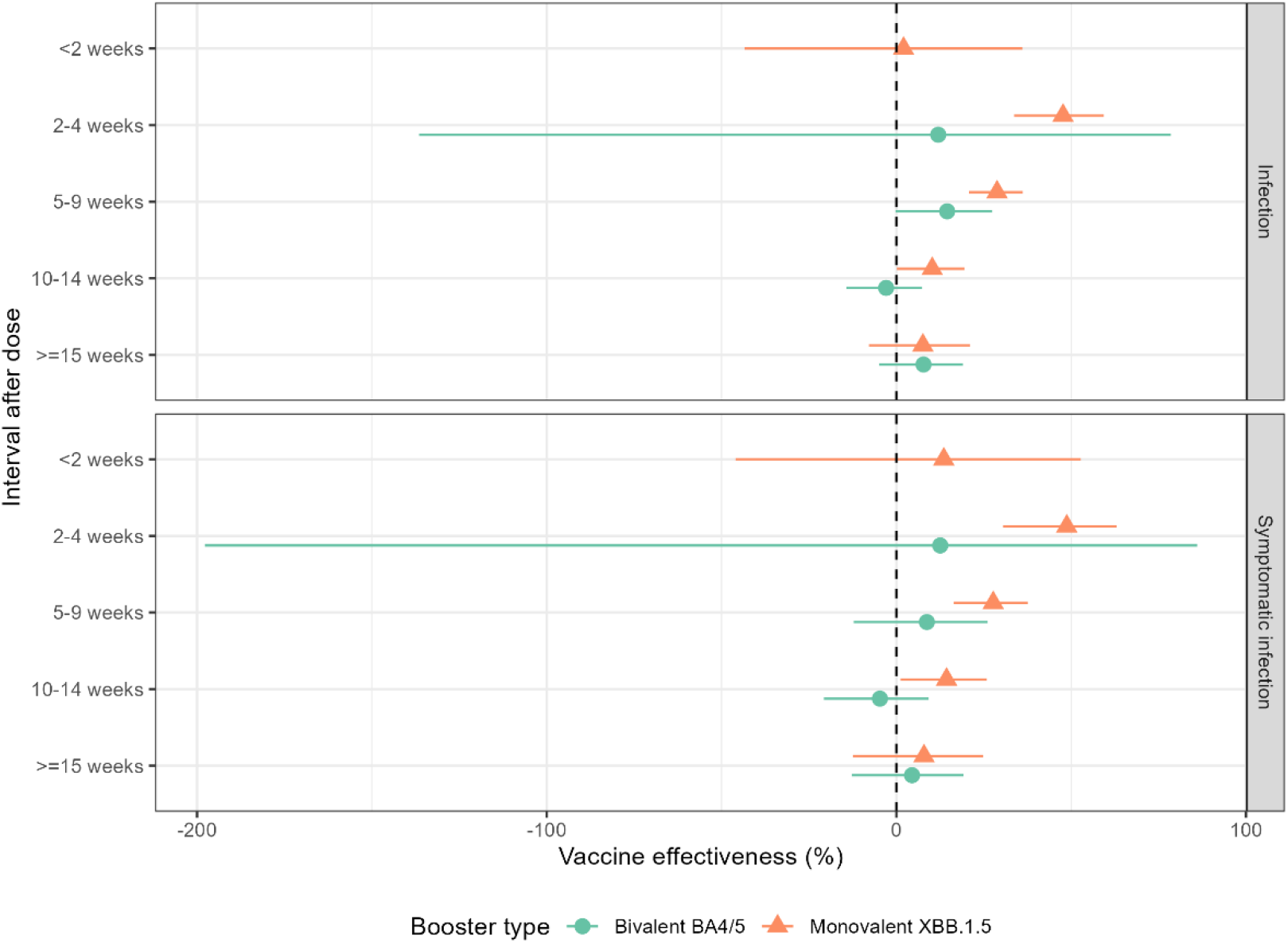
Vaccine effectiveness (VE) against infection and symptomatic infection amongst the WCIS study population, stratified by time since vaccination and vaccine type. Error bars represent 95% confidence intervals. Data behind estimates shown are presented in Supplementary Table S3.

Similar to the infection outcome, autumn 2023 VE against symptomatic infection peaked at 49% (95% CI: 32-63%) 2 to 4 weeks after vaccination and waned to 5% (95% CI: -7-16%) by 10 to 14 weeks (Figure 3). When stratified by vaccine type (Figure 4), VE against symptomatic infection of the monovalent XBB vaccine peaked at 49% (95% CI: 31-63%) at 2 to 4 weeks and waned to 7.9% (95% CI: -12-25%) by 15 or more weeks. The bivalent BA4/5 vaccine did not show significant VE against symptomatic infection at any time interval. This difference in VE between vaccines was significant with p<0.001.

For both outcomes, VE point estimates differed by less than 3% at 2 weeks and over with overlapping CIs when a comparator group of unboosted individuals with at least two prior doses was used. Similarly, sensitivity analyses removing negative tests from individuals with a contributing positive test, removing positive tests where the infection episode may have begun prior to the study, and removing adjustment for prior infection all separately showed point estimates differing by less than 3% at 2 weeks and over, with overlapping CIs.

## Discussion

Using a TNCC study among individuals participating in the WCIS household study, we find the England autumn 2023 COVID-19 vaccine programme provided significant protection against infection and symptomatic infection. VE peaked at around 50% against both infection and symptomatic infection, waning at 10 weeks or more after vaccination. This is comparable to findings from the UK SIREN healthcare worker study (18), which found VE against infection estimated at 40% (95% CI: 19-57%), noting that the different sample population and demographics of the SIREN study, which was skewed towards mainly females between 35 to 64 years old.

When stratified by vaccine type, the monovalent XBB effectiveness peaked at 49% against both infection and symptomatic infection. The bivalent BA.4/5 vaccine effectiveness could not be assessed in the period shortly after vaccination when VE may have been at its peak, and only showed significant VE 5-9 weeks post vaccination for infection at 16% (95% CI: 0.3-29%). While some studies have shown evidence of the bivalent providing some protection against infection and symptomatic infection in earlier time periods, and with differing lineage profiles (19,20), low or non-significant VE of the BA.4/5 vaccine against symptomatic infection in the winter 2023-2024 period has also been seen in the SIREN study (VE: 15%, 95% CI: -55-54%) and a population-based study in Singapore (VE: 8%, 95% CI: 5-12%)(21). Another factor that may explain VE differences by vaccine is that the study period coincided primarily with a period of BA.2.86 sub-lineage dominance, in particular JN.1 after replacement of the previous dominant XBB sub-lineages occurring at the start of the study (22). The BA.2.86 sub-lineages are potentially more similar to the XBB lineage used in the monovalent compared to the BA.4/5 lineage used in the bivalent vaccine. However, despite this potential difference in VE for infection, this difference was not seen in effectiveness against hospitalisation; the bivalent vaccine still conferred protection against more severe outcomes in the same time period, as evidenced by a population-level TNCC study in England where bivalent VE against hospitalisation peaked at 45% (95% CI: 36-53%) (23).

The TNCC study design was chosen to mimic and provide direct comparisons with previous studies looking at VE against symptomatic infection during the period of community testing (8,10). This TNCC design is complementary to a cohort approach as it is relatively rapid to implement and analyse and allows matching on exposure to a respiratory infection and health care access when assessing tests associated with ARI symptoms.

Understanding VE against less severe outcomes of COVID-19 such as infection and symptomatic infections within the wider population, as well as time periods conferring most protection, will help inform vaccine policy. In doing so, policies can address the health and economic burden caused by symptomatic illness, and help to reduce infections, which may lead to further transmission within the community and to more vulnerable sub-populations. While WCIS data could have been used to estimate VE against hospitalisations in the wider population, initial scoping of the data set showed that there were not enough COVID-19-associated hospitalisations among the WCIS cohort to provide power for an analysis.

A limitation of the study is that the demographic represented in the WCIS is skewed towards an older population who are less economically deprived which reduces the generalisability of results (Supplementary Table 1). Improvements for future study designs would aim to increase representativeness, for example through provision of financial incentives. The study also relied on the use of LFDs which, whilst having high sensitivity and specificity, are not the gold standard polymerase chain reaction (PCR) test (24), meaning, for a small number of results, there may be non-differential misclassification biasing VE towards the null. LFDs also do not allow lineage data through sequencing which would help inform the effectiveness of the vaccines against the circulating lineages more definitively, although the dominance of the BA.2.86 sub-lineages in the wider population at the time of the study do give a strong indication of the likely variant causing infection. The study was limited in estimating short term VE for the BA.4/5 vaccine as well as the long-term VE due to the ending of the WCIS study. While we have attempted to adjust for immunity derived from prior infections, most community infections would have been undocumented in SGSS since the end of widespread testing; however, in a sensitivity analysis, results were similar to the main analysis when adjustment for prior infection was removed, suggesting that this may not cause a large bias or impact in vaccine effectiveness. Similar results have been seen in previous studies (25).

In summary, this study showed moderate vaccine effectiveness of the boosters distributed during the autumn 2023 COVID-19 vaccination campaign against infection and symptomatic infection, waning after a short period of time, with significant differences between the two vaccines used. This emphasizes the continued need to understand optimal time periods for vaccine distribution for COVID-19, and the interaction of vaccine variant selection and coordination on vaccine effectiveness, especially throughout a background of changing lineage and disease profiles, to best provide the population with on-going protection against the disease.

## Data Availability

The individual-level nature of the data used risks individuals being identified, or being able to self-identify, if the data are released publicly. Requests for access to these non-publicly available data should be directed to UKHSA.

## Acknowledgments

The authors would like to thank the UKHSA Surveillance and Immunity team and ONS WCIS analysis team for their contributions.

## Notes

### Competing Interest Statement

The Immunisations Department in the UK Health Security Agency provides vaccine manufacturers (including Pfizer) with post-marketing surveillance reports about pneumococcal and meningococcal disease which the companies are required to submit to the UK Licensing authority in compliance with their Risk Management Strategy. A cost recovery charge is made for these reports.

### Funding Statement

This study was undertaken as part of UKHSA routine work to monitor COVID-19. No external funding was received.

### Author Declarations

UKHSA has legal permission, provided by Regulation 3 of The Health Service (Control of Patient Information) Regulations 2002 to process confidential patient information under Section 3(i) (a)-(c), 3(i)(d) (i) and (ii) and 3(iii) as part of its outbreak response activities. This study falls within the research activities approved by the UK Health Security Agency (UKHSA) Research Ethics and Governance Group.

## References

1. Dabrera G, Allen H, Zaidi A, Flannagan J, Twohig K, Thelwall S, et al. Assessment of mortality and hospital admissions associated with confirmed infection with SARS-CoV-2 Alpha variant: a matched cohort and time-to-event analysis, England, October to December 2020. Cog-Uk Consortium, editor. Euro surveillance : bulletin Europeen sur les maladies transmissibles = European communicable disease bulletin. 2022;27(20).

2. Twohig KA, Nyberg T, Zaidi A, Thelwall S, Sinnathamby MA, Aliabadi S, et al. Hospital admission and emergency care attendance risk for SARS-CoV-2 delta (B.1.617.2) compared with alpha (B.1.1.7) variants of concern: a cohort study. The Lancet Infectious Diseases. 2022 Jan;22(1):35–42.

3. Nyberg T, Twohig KA, Harris RJ, Seaman SR, Flannagan J, Allen H, et al. Risk of hospital admission for patients with SARS-CoV-2 variant B.1.1.7: cohort analysis. BMJ (Clinical research ed). 2021;373:n1412.

4. Stowe J, Andrews N, Kirsebom F, Ramsay M, Bernal JL. Effectiveness of COVID-19 vaccines against Omicron and Delta hospitalisation, a test negative case-control study. Nat Commun. 2022 Sep 30;13(1):5736.

5. Kirsebom FCM, Andrews N, Stowe J, Toffa S, Sachdeva R, Gallagher E, et al. COVID-19 vaccine effectiveness against the omicron (BA.2) variant in England. The Lancet Infectious Diseases. 2022 Jul;22(7):931–3.

6. Kirsebom FCM, Andrews N, Stowe J, Groves N, Chand M, Ramsay M, et al. Effectiveness of the COVID-19 vaccines against hospitalisation with Omicron sub-lineages BA.4 and BA.5 in England. The Lancet Regional Health - Europe. 2022 Dec;23:100537.

7. UK Health Security Agency. COVID-19: the green book, chapter 14a. Immunisation against infectious diseases. 2020.

8. Andrews N, Stowe J, Kirsebom F, Toffa S, Rickeard T, Gallagher E, et al. Covid-19 Vaccine Effectiveness against the Omicron (B.1.1.529) Variant. N Engl J Med. 2022 Apr 21;386(16):1532–46.

9. Andrews N, Tessier E, Stowe J, Gower C, Kirsebom F, Simmons R, et al. Duration of Protection against Mild and Severe Disease by Covid-19 Vaccines. N Engl J Med. 2022 Jan 27;386(4):340–50.

10. Andrews N, Stowe J, Kirsebom F, Toffa S, Sachdeva R, Gower C, et al. Effectiveness of COVID-19 booster vaccines against COVID-19-related symptoms, hospitalization and death in England. Nat Med. 2022 Apr;28(4):831–7.

11. UK Health Security Agency. GOV.UK. [cited 2022 Sep 29]. Changes to COVID-19 testing in England from 1 April. Available from: https://www.gov.uk/government/news/changes-to-covid-19-testing-in-england-from-1-april

12. Department of Health & Social Care. GOV.UK. [cited 2024 Dec 5]. JCVI statement on the COVID-19 vaccination programme for autumn 2023 - update 7 July 2023. Available from: https://www.gov.uk/government/publications/covid-19-autumn-2023-vaccination-programme-jcvi-update-7-july-2023/jcvi-statement-on-the-covid-19-vaccination-programme-for-autumn-2023-update-7-july-2023

13. Office for National Statistics. Office for National Statistics. [cited 2024 Dec 5]. Winter Coronavirus (COVID-19) Infection Study. Available from: https://www.ons.gov.uk/surveys/informationforhouseholdsandindividuals/householdandindividualsurveys/wintercoronaviruscovid19infectionstudy/aboutthewintercoronaviruscovid19infectionstudy

14. Office for National Statistics. Office for National Statistics. 2024 [cited 2024 Dec 15]. Winter Coronavirus (COVID-19) Infection Study QMI. Available from: https://www.ons.gov.uk/peoplepopulationandcommunity/healthandsocialcare/conditionsanddiseases/methodologies/wintercoronaviruscovid19infectionstudyqmi#quality-summary

15. Tessier E, Edelstein M, Tsang C, Kirsebom F, Gower C, Campbell CNJ, et al. Monitoring the COVID-19 immunisation programme through a national immunisation Management system – England’s experience. International Journal of Medical Informatics. 2023 Feb 1;170:104974.

16. NHS Digital. Cohorting as a Service (CaaS) [Internet]. 2023. Available from: https://digital.nhs.uk/services/cohorting-as-a-service-caas

17. Clare T, Twohig KA, O’Connell AM, Dabrera G. Timeliness and completeness of laboratory-based surveillance of COVID-19 cases in England. Public Health. 2021 Apr;S0033350621001219.

18. Kirwan PD, Foulkes S, Munro K, Sparkes D, Singh J, Henry A, et al. Protection of vaccine boosters and prior infection against mild/asymptomatic and moderate COVID-19 infection in the UK SIREN healthcare worker cohort: October 2023 to March 2024. J Infect. 2024 Nov;89(5):106293.

19. Rudolph AE, Khan FL, Shah A, Singh TG, Wiemken TL, Puzniak LA, et al. Effectiveness of BNT162b2 BA.4/5 Bivalent mRNA Vaccine Against Symptomatic COVID-19 Among Immunocompetent Individuals Testing at a Large US Retail Pharmacy. J Infect Dis. 2024 Mar 14;229(3):648–59.

20. Plumb ID, Briggs Hagen M, Wiegand R, Dumyati G, Myers C, Harland KK, et al. Effectiveness of a bivalent mRNA vaccine dose against symptomatic SARS-CoV-2 infection among U.S. Healthcare personnel, September 2022–May 2023. Vaccine. 2024 Apr 11;42(10):2543–52.

21. Chong C, Wee LE, Jin X, Zhang M, Malek MIA, Ong B, et al. Risks of SARS-CoV-2 JN.1 Infection and COVID-19-Associated Emergency Department Visits/Hospitalizations Following Updated Boosters and Prior Infection: A Population-Based Cohort Study. Clin Infect Dis. 2024 Nov 22;79(5):1190–6.

22. UK Health Security Agency. National Influenza and COVID-19 Report: week 13 report (up to week 12 2024 data) [Internet]. GOV.UK; 2024 Mar. Available from: https://assets.publishing.service.gov.uk/media/660546e0e8c442001122043a/Weekly-flu-and-COVID-19-surveillance-report_wk13.pdf.pdf

23. Kirsebom FCM, Stowe J, Lopez Bernal J, Allen A, Andrews N. Effectiveness of autumn 2023 COVID-19 vaccination and residual protection of prior doses against hospitalisation in England, estimated using a test-negative case-control study. Journal of Infection. 2024 Jul 1;89(1):106177.

24. Department of Health & Social Care, UK Health Security Agency. GOV.UK. 2023 [cited 2024 Dec 13]. Performance of lateral flow devices during the COVID-19 pandemic. Available from: https://www.gov.uk/government/publications/lateral-flow-device-performance-data/performance-of-lateral-flow-devices-during-the-covid-19-pandemic

25. Kirsebom FCM, Andrews N, Stowe J, Dabrera G, Ramsay M, Bernal JL. Effectiveness of the Sanofi/GSK (VidPrevtyn Beta) and Pfizer-BioNTech (Comirnaty Original/Omicron BA.4-5) bivalent vaccines against hospitalisation in England. eClinicalMedicine [Internet]. 2024 May 1 [cited 2024 Dec 5];71. Available from: https://www.thelancet.com/journals/eclinm/article/PIIS2589-5370(24)00166-4/fulltext

