## Supplementary Document for "Effectiveness of COVID-19 vaccines against SARS-CoV-2 infection and symptomatic infection in the England Winter Coronavirus (COVID-19) Infection Study cohort"

Nurin Abdul Aziz[1], Nick Andrews[1], Christopher Rawlinson[1], Andrew Buckley[2], Alexander Allen[1]

[1]Immunisations and Vaccine-Preventable Diseases Division, UK Health Security Agency

[2]Surveillance Strategy and Transformation Branch, UK Health Security Agency

**Supplementary Table 1**. Descriptive characteristics of cases and controls included in the analysis of the effectiveness of Autumn 2023 programme vaccines against infection and symptomatic infection among the Winter COVID-19 Study cohort

| | Infection outcome | | | | Symptomatic infection outcome | | | |
|---|---|---|---|---|---|---|---|---|
| | Controls | | Cases | | Controls | | Cases | |
| | n | % | n | % | n | % | n | % |
| **COVID Autumn 2023 vaccine status** | | | | | | | | |
| No Autumn 23 booster | 46,446 | 42.91 | 2,945 | 50.65 | 17,468 | 45.62 | 2,080 | 51.33 |
| Bivalent BA4/5 | 27,756 | 25.64 | 1,386 | 23.84 | 9,167 | 23.94 | 974 | 24.04 |
| Monovalent XBB | 33,888 | 31.30 | 1,476 | 25.39 | 11,594 | 30.28 | 993 | 24.51 |
| Other Autumn 23 | 142 | 0.13 | 6 | 0.10 | 50 | 0.13 | 4 | 0.10 |
| Autumn 23, missing vaccine type data | 4 | 0.00 | 0 | 0.00 | 3 | 0.01 | 0 | 0.00 |
| Unknown vaccination status | 16 | 0.01 | 1 | 0.02 | 6 | 0.02 | 1 | 0.02 |
| **Age group** | | | | | | | | |
| 0-9 | 766 | 0.71 | 33 | 0.57 | 351 | 0.92 | 28 | 0.69 |
| 10-19 | 3,156 | 2.92 | 124 | 2.13 | 1,112 | 2.90 | 83 | 2.05 |
| 20-29 | 1,360 | 1.26 | 69 | 1.19 | 470 | 1.23 | 53 | 1.31 |
| 30-39 | 5,475 | 5.06 | 390 | 6.71 | 2,270 | 5.93 | 272 | 6.71 |
| 40-49 | 12,084 | 11.16 | 879 | 15.12 | 4,805 | 12.55 | 610 | 15.05 |
| 50-59 | 21,632 | 19.98 | 1,337 | 23.00 | 8,173 | 21.35 | 949 | 23.42 |
| 60-69 | 31,046 | 28.68 | 1,569 | 26.99 | 11,143 | 29.10 | 1,089 | 26.88 |
| 70-79 | 27,598 | 25.49 | 1,239 | 21.31 | 8,529 | 22.28 | 845 | 20.85 |
| 80-89 | 4,916 | 4.54 | 165 | 2.84 | 1,378 | 3.60 | 117 | 2.89 |
| ≥90 | 219 | 0.20 | 9 | 0.15 | 57 | 0.15 | 6 | 0.15 |
| **Sex** | | | | | | | | |
| Female | 61,426 | 56.74 | 3,382 | 58.17 | 22,579 | 58.97 | 2,392 | 59.03 |
| Male | 46,826 | 43.26 | 2,432 | 41.83 | 15,709 | 41.03 | 1,660 | 40.97 |
| **Ethnic group** | | | | | | | | |
| Asian or Asian British | 2,412 | 2.23 | 100 | 1.72 | 659 | 1.72 | 67 | 1.65 |
| Black, Black British, Caribbean, or African | 362 | 0.33 | 19 | 0.33 | 119 | 0.31 | 13 | 0.32 |
| Mixed or multiple ethnic groups | 1,235 | 1.14 | 74 | 1.27 | 446 | 1.16 | 49 | 1.21 |
| White | 103,156 | 95.29 | 5,563 | 95.68 | 36,765 | 96.02 | 3,889 | 95.98 |
| Other ethnic group | 812 | 0.75 | 49 | 0.84 | 249 | 0.65 | 29 | 0.72 |
| Unknown | 275 | 0.25 | 9 | 0.15 | 50 | 0.13 | 5 | 0.12 |
| **IMD decile** | | | | | | | | |
| 1 | 3,196 | 2.95 | 142 | 2.44 | 1,216 | 3.18 | 102 | 2.52 |
| 2 | 4,882 | 4.51 | 249 | 4.28 | 1,825 | 4.77 | 175 | 4.32 |
| 3 | 6,704 | 6.19 | 347 | 5.97 | 2,418 | 6.32 | 248 | 6.12 |
| 4 | 8,767 | 8.10 | 458 | 7.88 | 3,140 | 8.20 | 308 | 7.60 |
| 5 | 10,632 | 9.82 | 516 | 8.88 | 3,766 | 9.84 | 353 | 8.71 |
| 6 | 12,404 | 11.46 | 678 | 11.66 | 4,421 | 11.55 | 493 | 12.17 |
| 7 | 13,665 | 12.62 | 762 | 13.11 | 4,776 | 12.47 | 514 | 12.69 |
| 8 | 14,774 | 13.65 | 805 | 13.85 | 5,211 | 13.61 | 563 | 13.89 |
| 9 | 15,509 | 14.33 | 880 | 15.14 | 5,382 | 14.06 | 641 | 15.82 |
| 10 | 17,719 | 16.37 | 977 | 16.80 | 6,133 | 16.02 | 655 | 16.16 |
| **Region** | | | | | | | | |
| East Midlands | 8,734 | 8.07 | 452 | 7.77 | 3,102 | 8.10 | 336 | 8.29 |
| East of England | 13,544 | 12.51 | 774 | 13.31 | 4,609 | 12.04 | 545 | 13.45 |
| London | 17,355 | 16.03 | 1,042 | 17.92 | 5,978 | 15.61 | 679 | 16.76 |
| North East | 4,896 | 4.52 | 222 | 3.82 | 1,851 | 4.83 | 160 | 3.95 |
| North West | 13,415 | 12.39 | 646 | 11.11 | 4,824 | 12.60 | 453 | 11.18 |
| South East | 18,183 | 16.80 | 1,048 | 18.03 | 6,416 | 16.76 | 722 | 17.82 |
| South West | 12,312 | 11.37 | 695 | 11.95 | 4,374 | 11.42 | 486 | 11.99 |

| | Infection outcome | | | | Symptomatic infection outcome | | | |
|---|---|---|---|---|---|---|---|---|
| | Controls | | Cases | | Controls | | Cases | |
| | n | % | n | % | n | % | n | % |
| West Midlands | 9,455 | 8.73 | 454 | 7.81 | 3,306 | 8.63 | 315 | 7.77 |
| Yorkshire and The Humber | 10,358 | 9.57 | 481 | 8.27 | 3,828 | 10.00 | 356 | 8.79 |
| **Healthcare/social care worker status** | | | | | | | | |
| Yes | 101,494 | 93.76 | 5,414 | 93.12 | 35,645 | 93.10 | 3,783 | 93.36 |
| No | 6,758 | 6.24 | 400 | 6.88 | 2,643 | 6.90 | 269 | 6.64 |
| **Flu vaccination (2023-2024) status** | | | | | | | | |
| Yes | 44,467 | 41.08 | 2,766 | 47.57 | 16,339 | 42.67 | 1,933 | 47.70 |
| No | 63,785 | 58.92 | 3,048 | 52.43 | 21,949 | 57.33 | 2,119 | 52.30 |
| **Prior infection status** | | | | | | | | |
| Yes | 99,542 | 91.95 | 5,426 | 93.33 | 34,937 | 91.25 | 3,798 | 93.73 |
| No | 8,710 | 8.05 | 388 | 6.67 | 3,351 | 8.75 | 254 | 6.27 |

**Supplementary Table 2.** Vaccine effectiveness (VE) against infection and symptomatic infection amongst the WCIS study population, stratified by time since vaccination

| Booster received | Interval after dose | Controls | | Cases | | **Vaccine effectiveness** (%) (95% CI) |
|---|---|---|---|---|---|---|
| | | n | % | n | % | |
| **Infection** | | | | | | |
| No booster | - | 46,446 | 42.91 | 2,945 | 50.65 | - |
| Autumn 23 booster | <2 weeks | 474 | 0.44 | 26 | 0.45 | 4.74 (-39.55-37.83) |
| | 2-4 weeks | 2,707 | 2.50 | 75 | 1.29 | 48.7 (34.95-60.13) |
| | 5-9 weeks | 16,679 | 15.41 | 741 | 12.75 | 27.64 (19.26-35.2) |
| | 10-14 weeks | 18,991 | 17.54 | 1,360 | 23.39 | 5.02 (-5.08-14.14) |
| | >=15 weeks | 22,939 | 21.19 | 666 | 11.46 | 8.44 (-3.77-19.24) |
| **Symptomatic infection** | | | | | | |
| No booster | - | 17,468 | 45.62 | 2,080 | 51.33 | - |
| Autumn 23 booster | <2 weeks | 145 | 0.38 | 14 | 0.35 | 16.77 (-40.45-54.49) |
| | 2-4 weeks | 858 | 2.24 | 48 | 1.18 | 49.24 (31.61-63.16) |
| | 5-9 weeks | 5,409 | 14.13 | 495 | 12.22 | 24.71 (13.85-34.27) |
| | 10-14 weeks | 7,430 | 19.41 | 943 | 23.27 | 5.35 (-7.09-16.35) |
| | >=15 weeks | 6,972 | 18.21 | 471 | 11.62 | 5.09 (-10.44-18.47) |

**Supplementary Table 3.** Vaccine effectiveness (VE) against infection and symptomatic infection amongst the WCIS study population, stratified by time since vaccination and vaccine type

| Booster received | Interval after dose | Controls n | Controls % | Cases n | Cases % | **Vaccine effectiveness** (%) (95% CI) |
|---|---|---|---|---|---|---|
| **Infection** | | | | | | |
| No booster | - | 46,446 | 42.97 | 2,945 | 50.71 | - |
| Bivalent BA4/5 | <2 weeks | 4 | 0.00 | 0 | 0.00 | - |
| | 2-4 weeks | 88 | 0.08 | 3 | 0.05 | 13.26 (-133.4-78.81) |
| | 5-9 weeks | 5,820 | 5.38 | 212 | 3.65 | 15.62 (0.25-28.87) |
| | 10-14 weeks | 8,810 | 8.15 | 756 | 13.02 | -1.49 (-13.96-9.64) |
| | >=15 weeks | 13,034 | 12.06 | 415 | 7.15 | 9 (-4.47-20.82) |
| Monovalent XBB | <2 weeks | 469 | 0.43 | 26 | 0.45 | 2.19 (-43.35-36.19) |
| | 2-4 weeks | 2,618 | 2.42 | 72 | 1.24 | 48.48 (34.39-60.16) |
| | 5-9 weeks | 10,828 | 10.02 | 529 | 9.11 | 30.01 (21.25-37.87) |
| | 10-14 weeks | 10,138 | 9.38 | 600 | 10.33 | 11.57 (0.57-21.41) |
| | >=15 weeks | 9,835 | 9.10 | 249 | 4.29 | 8.88 (-7.16-22.72) |
| **Symptomatic infection** | | | | | | |
| No booster | - | 17,468 | 45.69 | 2,080 | 51.40 | - |
| Bivalent BA4/5 | <2 weeks | 2 | 0.01 | 0 | 0.00 | - |
| | 2-4 weeks | 26 | 0.07 | 2 | 0.05 | 12.53 (-197.78-86.03) |
| | 5-9 weeks | 1,702 | 4.45 | 145 | 3.58 | 8.71 (-12.25-26.13) |
| | 10-14 weeks | 3,434 | 8.98 | 526 | 13.00 | -4.72 (-20.72-9.21) |
| | >=15 weeks | 4,003 | 10.47 | 301 | 7.44 | 4.47 (-12.79-19.21) |
| Monovalent XBB | <2 weeks | 143 | 0.37 | 14 | 0.35 | 13.56 (-46.02-52.76) |
| | 2-4 weeks | 832 | 2.18 | 46 | 1.14 | 48.72 (30.52-63.03) |
| | 5-9 weeks | 3,702 | 9.68 | 350 | 8.65 | 27.69 (16.36-37.58) |
| | 10-14 weeks | 3,982 | 10.42 | 415 | 10.25 | 14.35 (1.16-25.86) |
| | >=15 weeks | 2,935 | 7.68 | 168 | 4.15 | 7.9 (-12.43-24.84) |

**Supplementary Table 4**. Sensitivity analysis: VE against infection and symptomatic infection amongst the WCIS study population, with a comparator group of tests associated with no Autumn 2023 booster and at least 2 previous doses

| | Controls | | Cases | | |
|---|---|---|---|---|---|
| **Interval after dose** | n | % | n | % | **Vaccine effectiveness** (%) (95% CI) |
| **Infection** | | | | | |
| No booster + 2 doses | 44,249 | 40.88 | 2,842 | 48.88 | - |
| <2 weeks | 474 | 0.44 | 26 | 0.45 | 5.79 (-38.03-38.52) |
| 2-4 weeks | 2,707 | 2.50 | 75 | 1.29 | 49.27 (35.67-60.58) |
| 5-9 weeks | 16,679 | 15.41 | 741 | 12.75 | 28.47 (20.16-35.96) |
| 10-14 weeks | 18,991 | 17.54 | 1,360 | 23.39 | 5.8 (-4.27-14.9) |
| >=15 weeks | 22,939 | 21.19 | 666 | 11.46 | 7.01 (-5.5-18.06) |
| **Symptomatic infection** | | | | | |
| No booster + 2 doses | 16,666 | 43.53 | 1,997 | 49.28 | - |
| <2 weeks | 145 | 0.38 | 14 | 0.35 | 16.95 (-40.18-54.59) |
| 2-4 weeks | 858 | 2.24 | 48 | 1.18 | 49.56 (32.01-63.4) |
| 5-9 weeks | 5,409 | 14.13 | 495 | 12.22 | 25.32 (14.48-34.83) |
| 10-14 weeks | 7,430 | 19.41 | 943 | 23.27 | 5.62 (-6.87-16.64) |
| >=15 weeks | 6,972 | 18.21 | 471 | 11.62 | 2.99 (-13.06-16.78) |

**Supplementary Table 5**. Sensitivity analysis: VE against infection and symptomatic infection amongst the WCIS study population with negative tests removed from participants who had contributed a positive test

| | Controls | | Cases | | |
|---|---|---|---|---|---|
| **Interval after dose** | n | % | n | % | **Vaccine effectiveness** (%) (95% CI) |
| **Infection** | | | | | |
| No booster | 44,467 | 42.72 | 2,945 | 50.66 | - |
| <2 weeks | 441 | 0.42 | 26 | 0.45 | 6.92 (-36.47-39.29) |
| 2-4 weeks | 2,513 | 2.41 | 75 | 1.29 | 50.03 (36.62-61.18) |
| 5-9 weeks | 15,330 | 14.73 | 741 | 12.75 | 28.07 (19.72-35.59) |
| 10-14 weeks | 18,475 | 17.75 | 1,360 | 23.40 | 5.31 (-4.78-14.42) |
| >=15 weeks | 22,874 | 21.97 | 666 | 11.46 | 8.45 (-3.77-19.25) |
| **Symptomatic infection** | | | | | |
| No booster | 5,590 | 48.65 | 2,080 | 51.35 | - |
| <2 weeks | 41 | 0.36 | 14 | 0.35 | 19.44 (-36.02-55.97) |
| 2-4 weeks | 246 | 2.14 | 48 | 1.18 | 50 (32.6-63.72) |
| 5-9 weeks | 1,498 | 13.04 | 495 | 12.22 | 25.01 (14.16-34.55) |
| 10-14 weeks | 2,038 | 17.74 | 943 | 23.28 | 5.23 (-7.26-16.26) |
| >=15 weeks | 2,078 | 18.08 | 471 | 11.63 | 4.82 (-10.77-18.26) |

**Supplementary Table 6**. Sensitivity analysis: VE against infection and symptomatic infection amongst the WCIS study population with positive tests removed from participants where the COVID-19 episode may have begun prior to the WCIS study

| | Controls | | Cases | | |
|---|---|---|---|---|---|
| **Interval after dose** | n | % | n | % | **Vaccine effectiveness** (%) (95% CI) |
| **Infection** | | | | | |
| No booster | 46,446 | 42.91 | 2,902 | 51.23 | - |
| <2 weeks | 474 | 0.44 | 25 | 0.44 | 5.51 (-39.34-38.85) |
| 2-4 weeks | 2,707 | 2.50 | 73 | 1.29 | 48.59 (34.61-60.18) |
| 5-9 weeks | 16,679 | 15.41 | 710 | 12.53 | 28.82 (20.44-36.37) |
| 10-14 weeks | 18,991 | 17.55 | 1,303 | 23.00 | 7.29 (-2.72-16.32) |
| >=15 weeks | 22,939 | 21.19 | 652 | 11.51 | 9.23 (-3.02-20.04) |
| **Symptomatic infection** | | | | | |
| No booster | 5,894 | 48.90 | 2,053 | 52.03 | - |
| <2 weeks | 44 | 0.37 | 14 | 0.35 | 14.52 (-44.23-53.25) |
| 2-4 weeks | 270 | 2.24 | 46 | 1.17 | 50.22 (32.56-64.11) |
| 5-9 weeks | 1,646 | 13.66 | 472 | 11.96 | 26.58 (15.8-36.05) |
| 10-14 weeks | 2,116 | 17.56 | 902 | 22.86 | 7.84 (-4.45-18.69) |
| >=15 weeks | 2,082 | 17.28 | 459 | 11.63 | 6.11 (-9.44-19.48) |

**Supplementary Table 5**. Sensitivity analysis: VE against infection and symptomatic infection amongst the WCIS study population without adjustment for recent COVID-19 infections prior to the study

| **Interval after dose** | **Vaccine effectiveness** (%) (95% CI) |
|---|---|
| **Infection** | |
| No booster | - |
| <2 weeks | 4.61 (-39.74-37.74) |
| 2-4 weeks | 48.67 (34.92-60.11) |
| 5-9 weeks | 27.67 (19.29-35.22) |
| 10-14 weeks | 5.18 (-4.9-14.29) |
| >=15 weeks | 8.61 (-3.57-19.39) |
| **Symptomatic infection** | |
| No booster | - |
| <2 weeks | 15.92 (-41.85-54.01) |
| 2-4 weeks | 49.34 (31.74-63.23) |
| 5-9 weeks | 24.79 (13.94-34.34) |
| 10-14 weeks | 5.69 (-6.7-16.64) |
| >=15 weeks | 5.52 (-9.94-18.83) |
